# General Anesthesia vs Procedural Sedation and Post-Thrombectomy Stress Hyperglycemia: A Retrospective Cohort Study

**DOI:** 10.64898/2026.09.02.26362114

**Authors:** Ankith Laichetty, Keri Zhou, William C. Broaddus

**Affiliations:** Wake Forest University School of Medicine; Virginia Commonwealth University School of Medicine

**Keywords:** Acute ischemic stroke, stress hyperglycemia, perioperative care

## Abstract

**Background:** The optimal anesthetic strategy for mechanical thrombectomy remains contested; clinical trials and meta-analyses yield conflicting results. Emerging evidence has shown stress hyperglycemia ratio (SHR) to be among the strongest independent predictors of poor thrombectomy outcomes. General anesthesia (GA) is known to provoke a more pronounced neuroendocrine stress response than procedural sedation, yet no prior study has investigated perioperative glucose dynamics between GA and sedation in thrombectomy patients.

**Methods:** Using the TriNetX federated research network, we conducted a retrospective propensity score-matched cohort study of non-diabetic adults undergoing mechanical thrombectomy for anterior circulation acute ischemic stroke under GA versus sedation. After propensity score matching 2,556 matched pairs remained.

**Results:** At the population level, mean glucose divergence was modest in absolute terms (Day 0: 110.90 vs 106.77 mg/dL; Δ = 4.13 mg/dL; p = 7.82 x 10^-11^; Day 1: 107.69 vs 103.25 mg/dL; Δ = 4.44 mg/dL; p = 8.16 x 10^-14^). GA conferred an 83.2% relative risk increase to severe-range hyperglycemia, defined as blood glucose ≥ 181 mg/dL on the day of the index procedure (Day 0: RR = 1.832, 95% CI 1.457 – 2.303; NNH: 29), with effects persisting on day 1 following the thrombectomy (Day 1: RR = 1.66, 95% CI 1.228 – 2.249). GA patients received significantly more exogenous insulin on both days 0 (GA: 21.24%, Sedation: 17.23%, RR = 1.231, p = 2.96 x 10^-4^) and day 1 (GA: 11.3%, Sedation: 8.69%, RR = 1.30, p = 0.00178). All primary outcomes survived Bonferroni correction.

**Conclusion:** General anesthesia is strongly associated with persistent hyperglycemia following mechanical thrombectomy in non-diabetic patients. These findings identify anesthesia-modality as a potential modifiable risk and provides a mechanistic hypothesis in the ongoing GA versus CS debate for thrombectomy. Future glucose management protocols may need to be tailored to anesthetic strategy.

## Introduction

Thrombectomy is the definitive treatment for large vessel occlusions in the brain. Every minute of delay in re-perfusing brain tissue has a massive cost. In addition, there are conflicting results in multiple anesthesia trials regarding whether general anesthesia (GA) versus procedural sedation for thrombectomy leads to better outcomes. For instance, a 2019 JAMA meta-analysis found no significant difference between sedation and GA, but a 2026 pooled analysis found GA was associated with poorer functional outcomes, whereas the 2025 SEGA trial found GA to be associated with superior functional outcomes.^1–4^ There is significant equipoise within the literature, existing anesthesia research emphasizes hemodynamics as the mechanism for the difference in outcomes, but this does not sufficiently explain the ambiguity and conflicting results in several high-profile trials and meta-analyses.^2,5–15^

The role glucose plays in stroke is also unclear; the previous 2019 SHINE trial found that there was no benefit between intensive glucose control (target blood glucose <110 mg/dL) and standard glucose control group (target <180 mg/dL) for patients following thrombectomy for acute ischemic stroke (AIS).^16^ However, the 2019 HERMES meta-analysis found that glucose may play a substantial role in thrombectomy efficacy. Patients with glucose <90 mg/dL had an adjusted common odds ratio (acOR) of 3.81for functional outcomes compared to the >90 mg/dL group who had an acOR of 1.83 for functional outcomes; which implies that the lower glucose group had twice the treatment effect as the higher group.^4^ These results suggest that even minor differences in glucose (e.g., 20-30 mg/dL) may have a more significant impact on functional outcomes than previously considered. Anesthetic modality has been established to affect glucose; particularly the stress hyperglycemia response. Specifically, GA causes a more profound stress response than neuraxial anesthesia or procedural sedation techniques.^17^

Therein lies a paradox, procedural sedation causes less significant metabolic derangements, hyperglycemia worsens stroke outcomes, whereas there is little consensus among high power meta-analyses. While there is data for perioperative glucose dynamics, it is limited in that no randomized controlled trial (RCT) specifically investigates glucose dynamics in non-diabetic patients with sufficient power. Moreover, the brain is highly glucose dependent and relies almost exclusively on glucose for fuel, consuming 20% of the body’s glucose; making it uniquely sensitive to glucose dysregulation. In addition, there is overwhelming mechanistic evidence that hyperglycemia worsens ischemic reperfusion injury.^18–20^

There are parallels between thrombectomy in the setting of ischemic stroke and CABG procedures in the setting of myocardial infarction. Both operations re-perfuse an ischemic organ. Existing CABG guidelines emphasize that hyperglycemia worsens outcomes when re-perfusing the heart, and stringent protocols to correct dysglycemia are laid forth.^21,22^ Re-perfusing the brain is philosophically similar, and hyperglycemia may play a larger role than previously considered, as the brain is more glucose dependent than the heart and less tolerant of ischemia. Emerging evidence has placed stress hyperglycemia, quantified by the stress hyperglycemia ratio (SHR) as one of the strongest independent predictors of poor overall outcomes; and the effect of GA on the perioperative stress response in non-diabetic patients following mechanical thrombectomy for acute ischemic stroke is not represented in the literature.^23–25^

## Methods

### Setting

The TriNetX database is a global health-collaborative clinical-research platform collecting real-time medical data from a network of HCOs holds the largest global patient dataset. In this study we used the Research Collaborative Network in TriNetX to build a cohort out of the more than 118 HCOs, 161 million participants between 1 July 2006 and 1 August 2026. Because the network is continuously refreshed and the platform re-derives cohorts at each execution, all queries were run on 07/31/2026 and 08/01/2026, and all counts are reported as of that execution (complete definitions provided in Supplemental Methods 1 and full TriNetX query architecture is given in Supplemental Methods 3).

### Ethics Statement

The TriNetX platform is compliant with the Health Insurance Portability & Accountability Act and General Data Protection Regulation. The Western Institutional Review Board has granted TriNetX a waiver of informed consent since this platform only aggregated counts and statistical summaries of de-identified information.

### Cohort Construction

#### Index event

The Index event was derived from the cohort definitions and specified separately for each cohort; the index-event logic was identical to the full cohort-defining logic in each case, excluding the cohort-specific Neuromuscular blockading agent (NMBA) criteria. The index date was therefore the date of the qualifying mechanical thrombectomy at which all inclusion criteria were satisfied, and no exclusion criterion applied and was designated day 0 for all time-anchored analyses. Patients without a qualifying index event were ineligible.

#### Exposure definition

While anesthetic modality is directly coded in the network, it is not reliable and returns spuriously low patient counts. As such assignment was operationalized using NMBA exposure as a surrogate for endotracheal intubation and general anesthesia.^26^

#### Cohort 1 – General anesthesia (GA)

Patients with an NMBA administration recorded on the same calendar date as the thrombectomy.

#### Cohort 2 – Sedation

Patients meeting all shared criteria with no NMBA administration recorded within 2 days on or after the thrombectomy (day 0 through day +2).

Inclusion Criteria:

- Age: At least 18 years at the most recent recorded age.
- Index: Mechanical thrombectomy
- Anterior-circulation acute ischemic stroke
- Non-Diabetic baseline glycemic status: At least one hemoglobin A1c value of 6.40% or less (EHR-sourced) recorded 3 months before through day 1 after the thrombectomy.^27^

Exclusion Criteria:

- Coded stroke severity outside the target range or impaired consciousness, on the day of thrombectomy
- Same-day intracranial hemorrhage, periprocedural infarction, or hypertensive crisis
- Comorbidities recorded on or before the thrombectomy
- Posterior-circulation and venous infarction
- Pregnancy

### Pre-specified Outcomes

All outcomes were ascertained from coded EHR data within windows anchored to the index thrombectomy (day 0). Serum glucose values and insulin administration were assessed as discrete daily windows on days 0 and 1 relative to index. Analyses used the platform’s Measures of Association, Number of Instances, and Lab Result Distribution functions. Propensity score matching Cohorts were balanced by 1:1 propensity score matching using greedy nearest-neighbor matching with a caliper of 0.1 pooled standard deviations of the logit of the propensity score, with randomized record ordering. Balance was assessed by standardized mean differences, with values below 0.1 considered acceptable.

Endpoints for which no qualifying events were observed in either cohort are reported as zero-count outcomes; additionally, the network applies small-cell obfuscation to counts below 10, and any such counts are reported accordingly.

### Glycemic Outcomes Definitions

Glycemic outcomes were defined as the proportion of patients in each cohort falling in selected blood glucose ranges. Glucose thresholds were specified as the most-recent lab value in the selected time window. Glucose data was queried as a consolidated data point from either measure of absolute serum, plasma, or blood glucose concentrations.

- Clinically Significant Hypoglycemia: <60 mg/dL: Hypoglycemia^9,28^
- Moderate Target-Range Hyperglycemia:141-180 mg/dL^9,28^
- Severe Hyperglycemia: ≥181 mg/dL^9,29^

### Exogenous Insulin Administration

- Any instance of exogenous insulin administration

### Glucose Monitoring Intensity

- Point-Of-Care Glucose
- Quantitative Laboratory Glucose
- Composite Monitoring: Any instance of either POC Glucose, Quantitative Lab Glucose or Comprehensive Metabolic Panel

#### Propensity Score Matching

We adopted TriNetX built-in function and matched the two groups 1:1 ratio by greedy nearest neighbor matching up to index date index; for demographic factors, lifestyle and vascular risk factors, comorbidities, stroke location and mechanism, intravenous alteplase administration, stroke severity, lab value availability, and vital signs. Standardized difference (Std diff) was used to evaluate the balance of baseline characteristics in score-matched populations. Generally, Std diff <0.10 is considered a small difference (complete covariate specifications and definitions are detailed in Table 1 and Supplemental Methods 1).^30^

**Table 1.**
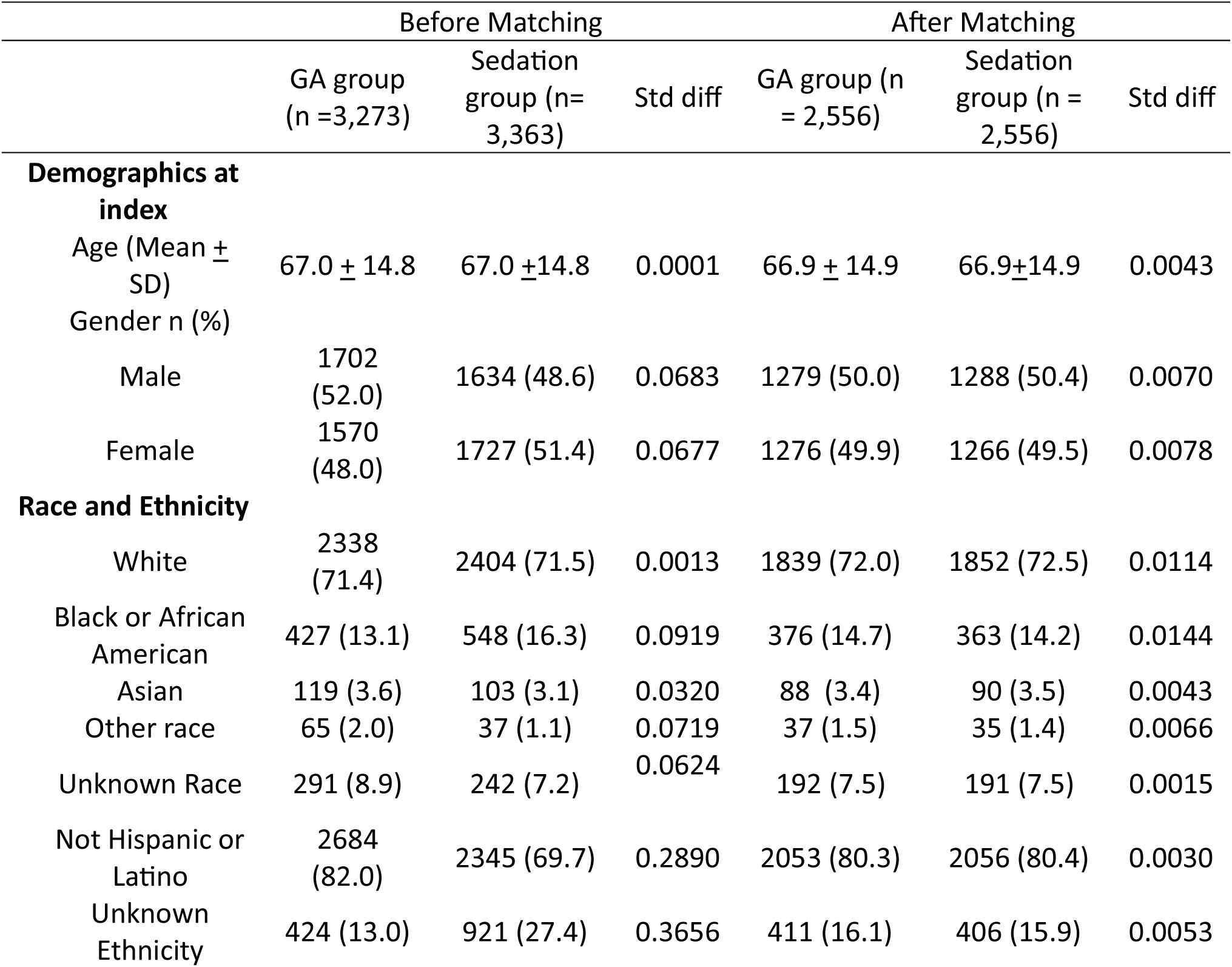
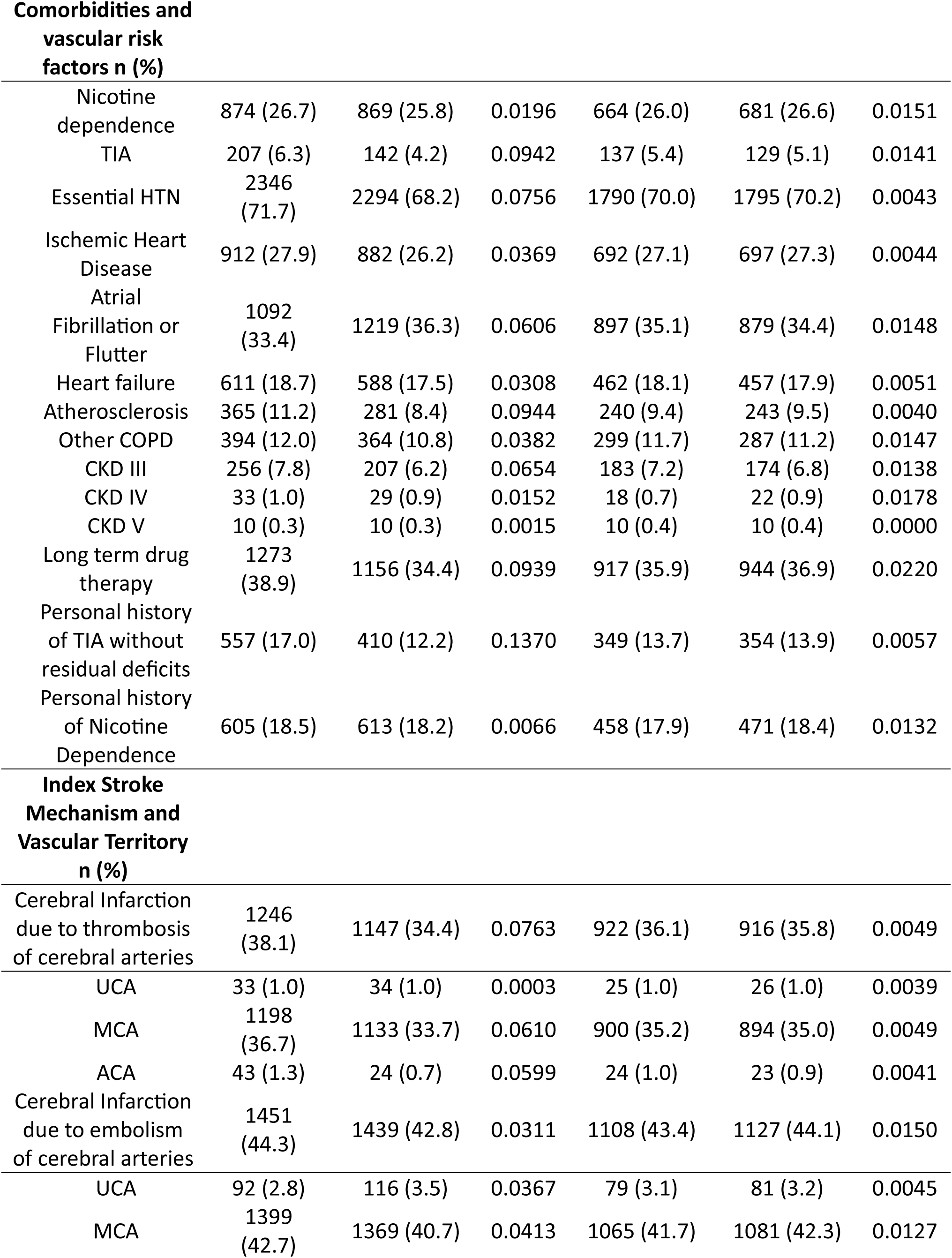
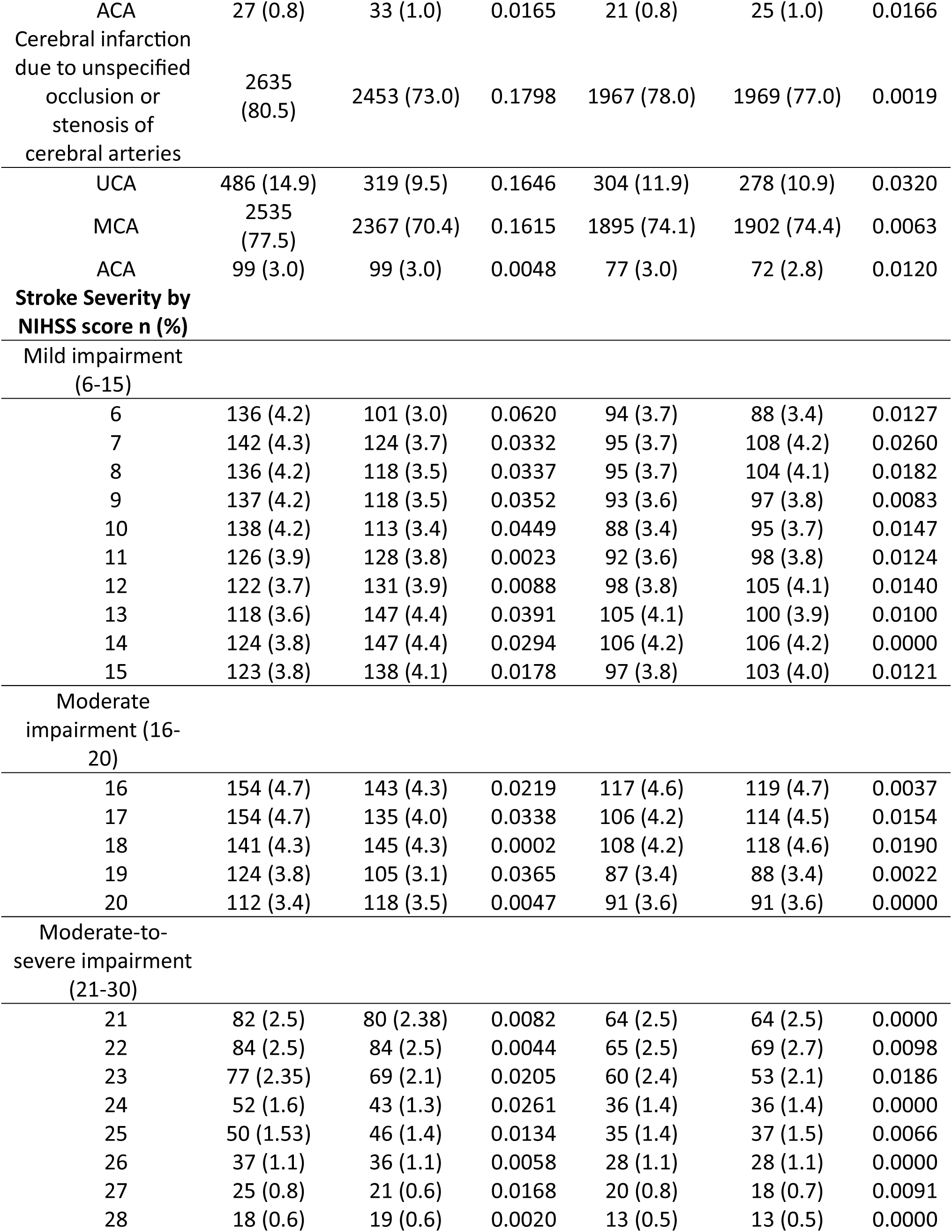
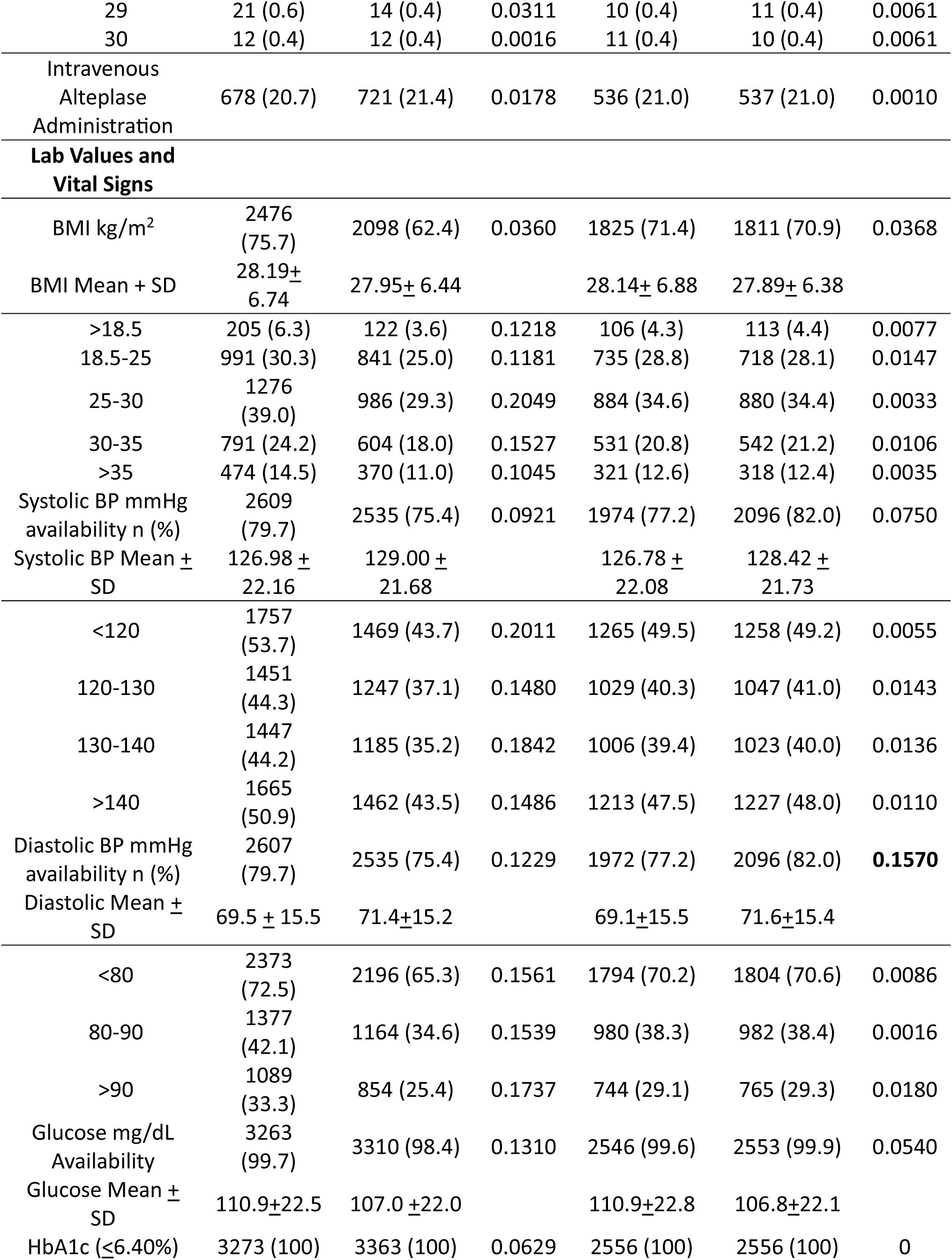
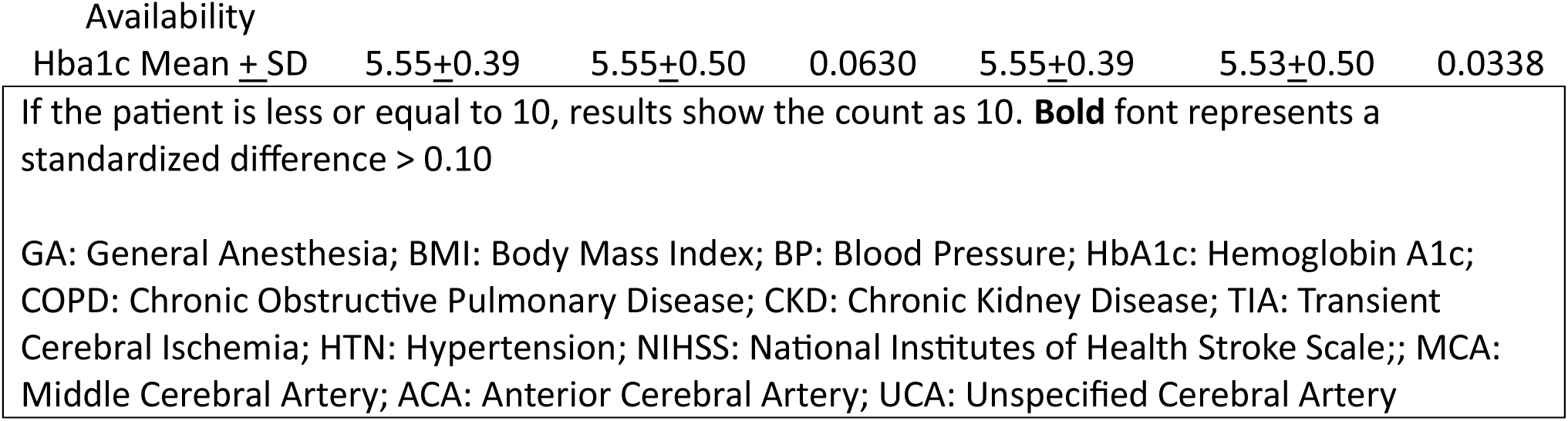
Baseline characteristics of study subjects (before and after Propensity score matching)

#### Statistical Analyses

For the pre-specified outcomes, measure of association analysis, patient counts with outcome, and distribution data was queried from the platform. In addition, T-Test statistics testing for the difference between the cohorts was performed. In all analyses, a 95% CI (95% CI) was considered evidence of statistical significance. Statistical significance was additionally defined as p-value < 0.05.

Sensitivity analysis was conducted using E-value point estimates for the Risk Ratio (RR) and the lower bound of the 95% CI of the RR.^31^ Bonferroni correction to be applied to all primary outcomes on both Day 0 and 1: Target-range Hyperglycemia, Severe Hyperglycemia, and the Insulin Administration outcomes; for deliberately conservative analysis.

## Results

From an initial network population of 30,978 patients undergoing mechanical thrombectomy, 6,800 met all eligibility criteria and were included in the propensity score matching analysis (figure 7). The database query returned 2,556 matched pairs (n = 5,112) after propensity score matching. Propensity score matching achieved excellent covariate balance across all matched parameters; Std diff remained well below 0.10 across all evaluated strata (maximum stratum Std diff = 0.0320 for unspecified cerebral artery occlusion; maximum diastolic blood pressure stratum Std diff = 0.0180; Table 1).

### Primary Outcomes

#### Perioperative glucose distribution statistics

Patients managed under GA demonstrated statistically significant increases in mean blood glucose compared to those receiving procedural sedation on both Day 0 and Day 1. Median glucose values were correspondingly higher in the GA cohort on both days (Day 0; GA:107 mg/dL, IQR = 26 vs. Sedation: 103 mg/dL, IQR = 24; Day 1; GA: 105 mg/dL, IQR = 24 vs. Sedation: 101 mg/dL, IQR = 21). **Day 0** (mean_GA_= 110.90±22.58 mg/dl vs mean_Sedation_ 106.77 ± 22.51 mg/dL; t = 6.52, p = 7.82 x 10^-11^). Among patients with available laboratory data in the immediate post procedural window (GA n = 2,512, Sedation n = 2,499). **Day 1** (mean_GA_ = 107.69±20.83 mg/dl vs mean_Sedation_ 103.25 ± 19.28 mg/dL; t = 7.49, p = 8.16 x 10^-14^). Among patients with available laboratory data in the immediate post procedural window (GA n = 2,349, Sedation n = 2,258) (Figures 1 and 4).

**Figure 1.**
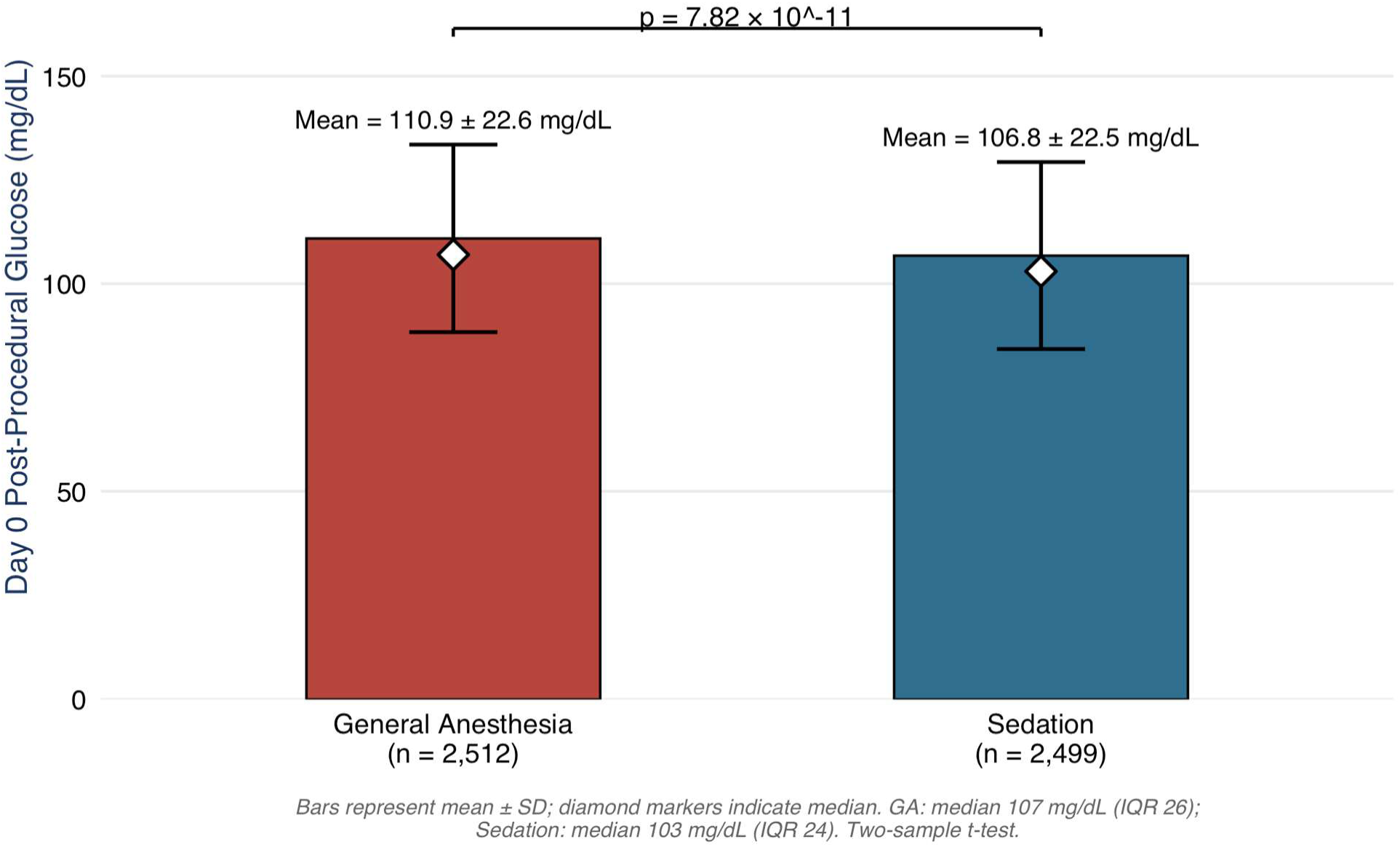
Day 0 Perioperative Glucose Distribution by Anesthetic Modality Mean post-procedural glucose on the day of index for mechanical thrombectomy (Day 0) in patients managed under general anesthesia (GA, n = 2512) versus procedural sedation (Sedation, n=2,499). Bars represent mean ± SD; diamond markers indicate median (GA: 107 mg/dL, IQR 26; Sedation: 103 mg/dL, IQR 24). GA was associated with significantly higher mean Day 0 glucose than Sedation (110.90 ± 22.58 vs. 106.77 ± 22.51 mg/dL; two-sample t-test, p = 7.82 × 10⁻¹¹).

#### Target-range hyperglycemia

GA patients experienced significantly higher risk of target-range hyperglycemia (141-180 mg/dL) on both days. **Day 0:** with Nearly a third of GA patients had this outcome (30.99%) vs 21.17% of the sedation cohort (GA: 30.99%[792/2,556] vs Sedation: 21.17%[541/2,556], z = 7.996, p = 1.29 x 10^-15^) with a relative increase in risk of 46.4% (RR = 1.464, 95% CI 1.332 – 1.610). **Day 1:** GA patients were again more represented in this tier with 18.3% having this outcome vs 13.3% of the Sedation cohort (GA: 18.3% [468/2,556] vs Sedation: 13.3% [340/2,556], z = 4.908, p = 9.22 x 10^-7^) with a relative increase in risk of 37.6% (RR = 1.376, 95% CI 1.211-1.565) (Figures 2 and 5).

**Figure 2.**
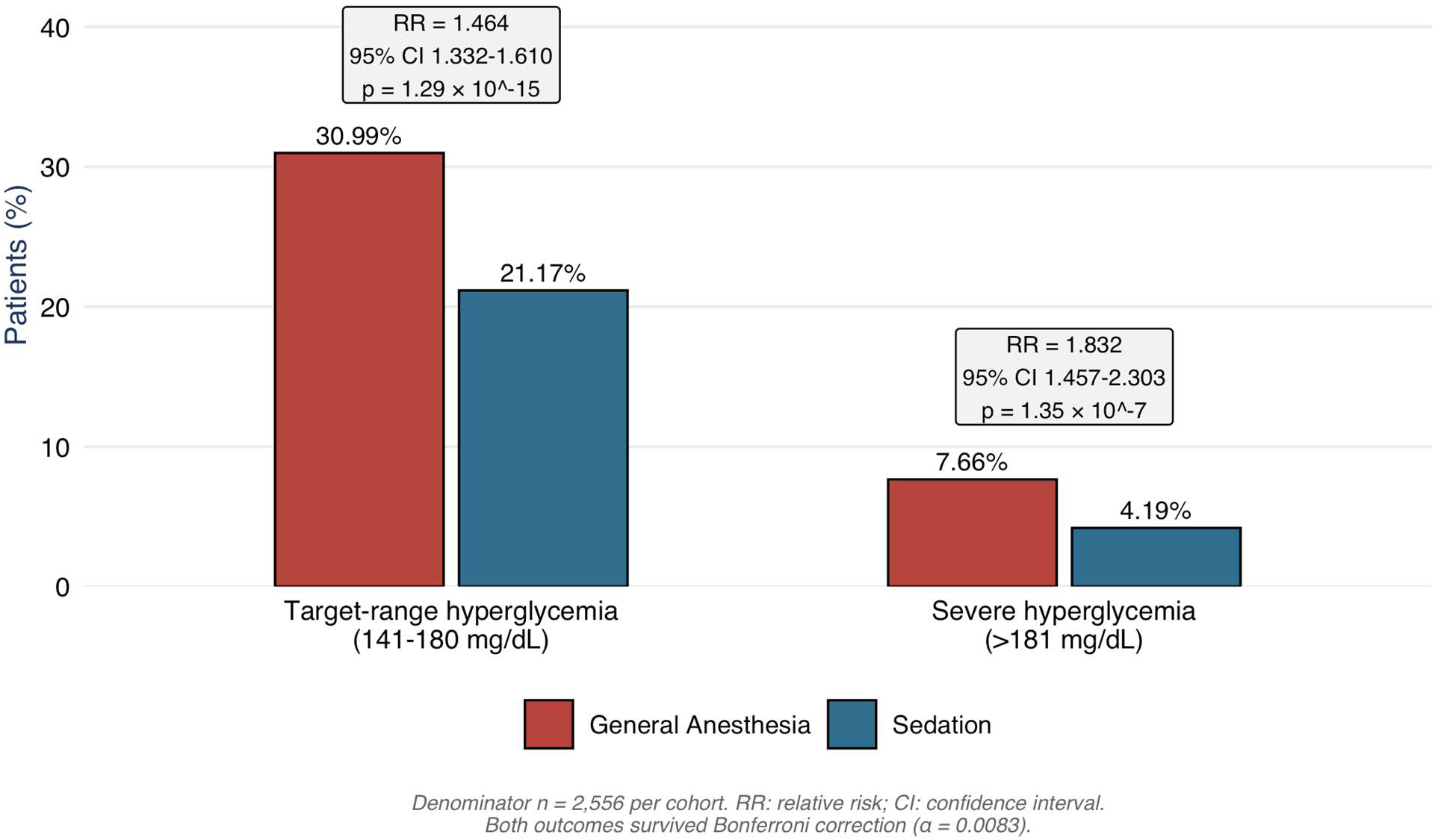
Day 0 Hyperglycemia Risk Stratification by Anesthetic Modality Proportion of patients meeting target-range hyperglycemia (141-180mg/dL) and severe hyperglycemia (>181 mg/dL) thresholds on day of index for mechanical thrombectomy (Day 0), by anesthetic modality (n=2,556 per cohort, post-propensity-score matching). GA patients were significantly more likely than Sedation patients to meet both thresholds; target-range hyperglycemia (30.99% vs. 21.17%; RR = 1.464, 95% CI 1.332–1.610; p = 1.29 × 10⁻¹⁵) and severe hyperglycemia (7.66% vs. 4.19%; RR = 1.832, 95% CI 1.457–2.303; p = 1.35 × 10⁻⁷). Both outcomes survived Bonferroni correction (α = 0.0083, six-outcome family)

**Figure 3.**
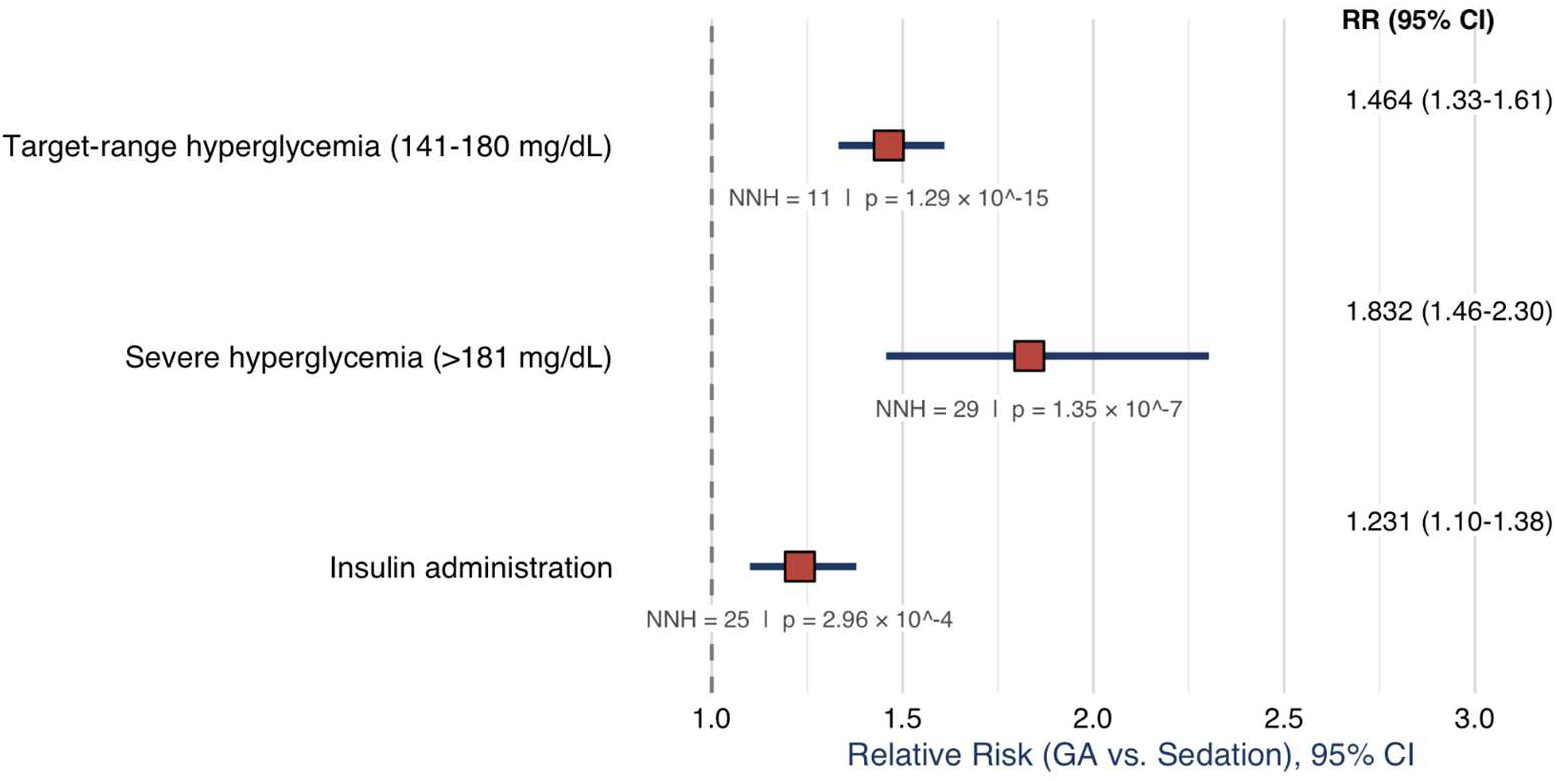
Forest Plot of Day 0 primary Glycemic outcomes Relative Risk (RR) of insulin administration, severe hyperglycemia (>181 mg/dL), and target range hyperglycemia (141-180 mg/dL) in patients managed under general anesthesia versus procedural sedation on Day 0. Squares represent point RR estimates; horizontal bars illustrate 95% confidence intervals; the dashed vertical line marks the null value (RR=1.0). All three outcomes favored increased risk under GA and survived Bonferroni Correction. NNH: number needed to harm.

**Figure 4.**
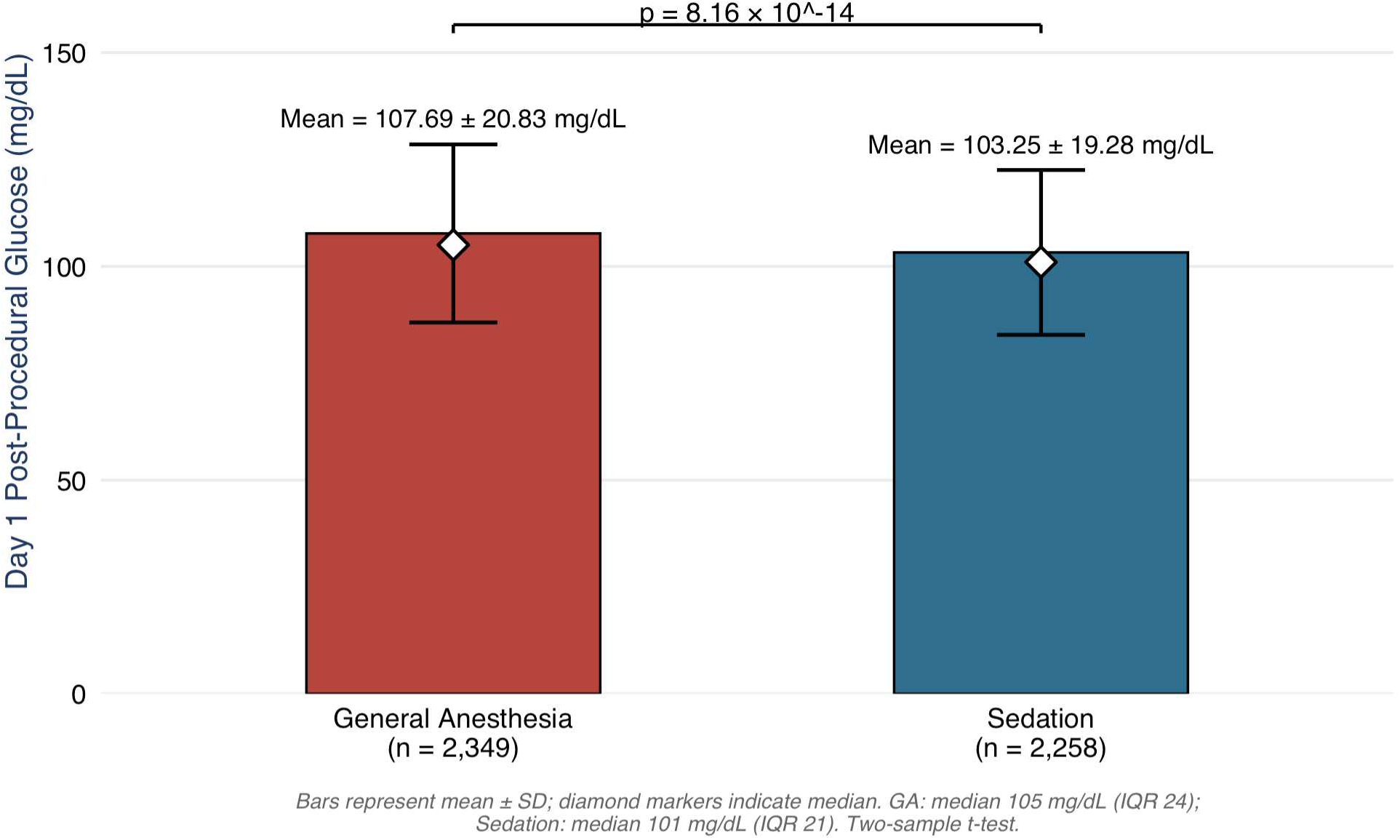
Day 1 Perioperative Glucose Distribution by Anesthetic Modality Mean post-procedural glucose on the day of index for mechanical thrombectomy (Day 0) in patients managed under general anesthesia (GA, n = 2349) versus procedural sedation (Sedation, n=2,258). Bars represent mean ± SD; diamond markers indicate median (GA: 105 mg/dL, IQR 24; Sedation: 101 mg/dL, IQR 21). GA was associated with significantly higher mean Day 1 glucose than Sedation (107.69 ± 20.83 vs. 103.25 ± 19.28 mg/dL; two-sample t-test, p = 8.16 × 10^-14^).

**Figure 5.**
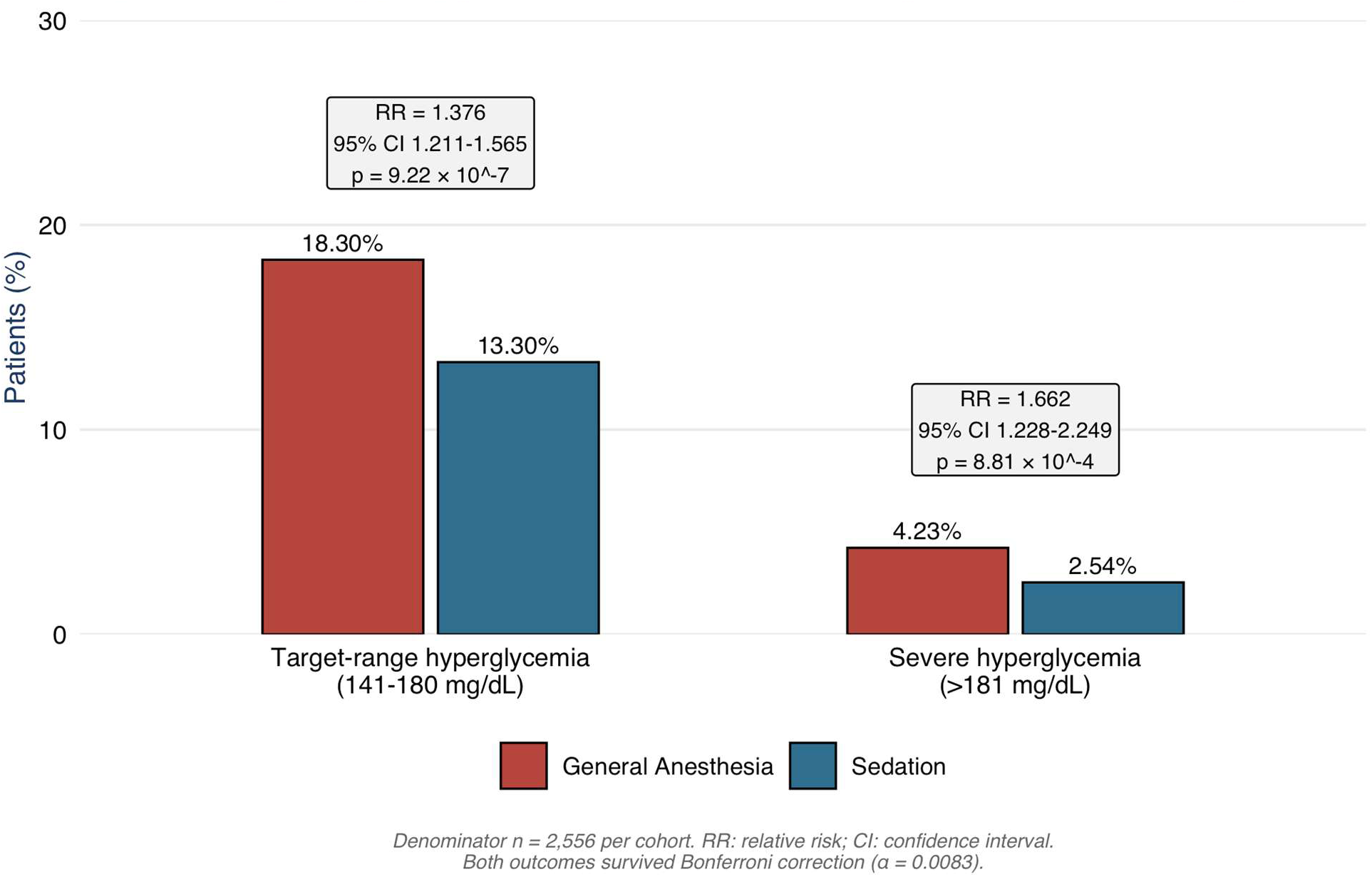
Day 1 Hyperglycemia Risk Stratification by Anesthetic Modality Proportion of patients meeting target-range hyperglycemia (141-180mg/dL) and severe hyperglycemia (>181 mg/dL) thresholds on Day 1 by anesthetic modality (n=2,556 per cohort, after propensity-score matching). GA patients were significantly more likely than Sedation patients to meet both thresholds; target-range hyperglycemia (18.3% vs. 13.3%; RR = 1.376, 95% CI 1.211–1.565; p =9.22 x 10^-7^) and severe hyperglycemia (4.23% vs. 2.54%; RR = 1.662, 95% CI 1.228–2.249; p = 8.81 x 10^-4^) Both outcomes survived Bonferroni correction (α = 0.0083, six-outcome family)

#### Severe hyperglycemia

GA patients experienced significantly higher risk of severe hyperglycemia (≥181 mg/dL) on both days. **Day 0:** Patients managed under GA demonstrated an 83.2% relative increase in severe same-day hyperglycemia compared to those managed under procedural sedation (GA: 7.66% [196/2,556] vs. Sedation: 4.19%[107/2,556]; RR = 1.832, 95% CI 1.457 – 2.303). This corresponded to a statistically significant absolute risk difference of 3.48% (95% CI 2.20% – 4.77%; z = 5.271, p = 1.35 x 10^-7^) and an odds ratio of 1.901 (95% CI: 1.492-2.422). **Day 1:** Patients managed under GA demonstrated a 66.2% relative increase in severe hyperglycemia compared to those managed under procedural sedation (GA: 4.23% [108/2,556] vs Sedation: 2.54% [65/2,556]; RR = 1.66, 95% CI 1.228 – 2.249). This corresponds to a statistically significant absolute risk difference of 1.68% (95% CI 0.692% vs. 2.67%, z = 3.326, p = 8.81 x 10^-4^) (Figures 2 and 5).

#### Exogenous Insulin Administration

Consequently, GA patients required significantly higher rates of exogenous insulin administration on both days compared to those in the Sedation cohort. **Day 0:** 21.24% (543/2,556) of GA patients received insulin therapy vs. 17.23% (441/2,556) of patients in the Sedation cohort. This represents an absolute risk increase of 3.99% (95% CI: 1.83%-6.15%; z = 3.618, p = 2.96 x 10^-4^), a 23.1% relative risk increase (RR = 1.231, 95% CI: 1.100-1.379), and an odds ratio of 1.294 (95% CI: 1.125 – 1.488). **Day 1:** 11.3% (289/2,556) of GA patients received insulin therapy vs. 8.69% (222/2,556) of patients in the Sedation cohort. This represents an absolute risk increase of 2.62%(95% CI 0.978% – 4.264%, z = 3.124, p = 0.00178) a 30% relative risk increase (RR = 1.30, 95% CI: 1.102 – 1.537), and an Odds Ratio of 1.340 (95% CI 1.11 – 1.61) (Figures 3 and 6).

**Figure 6.**
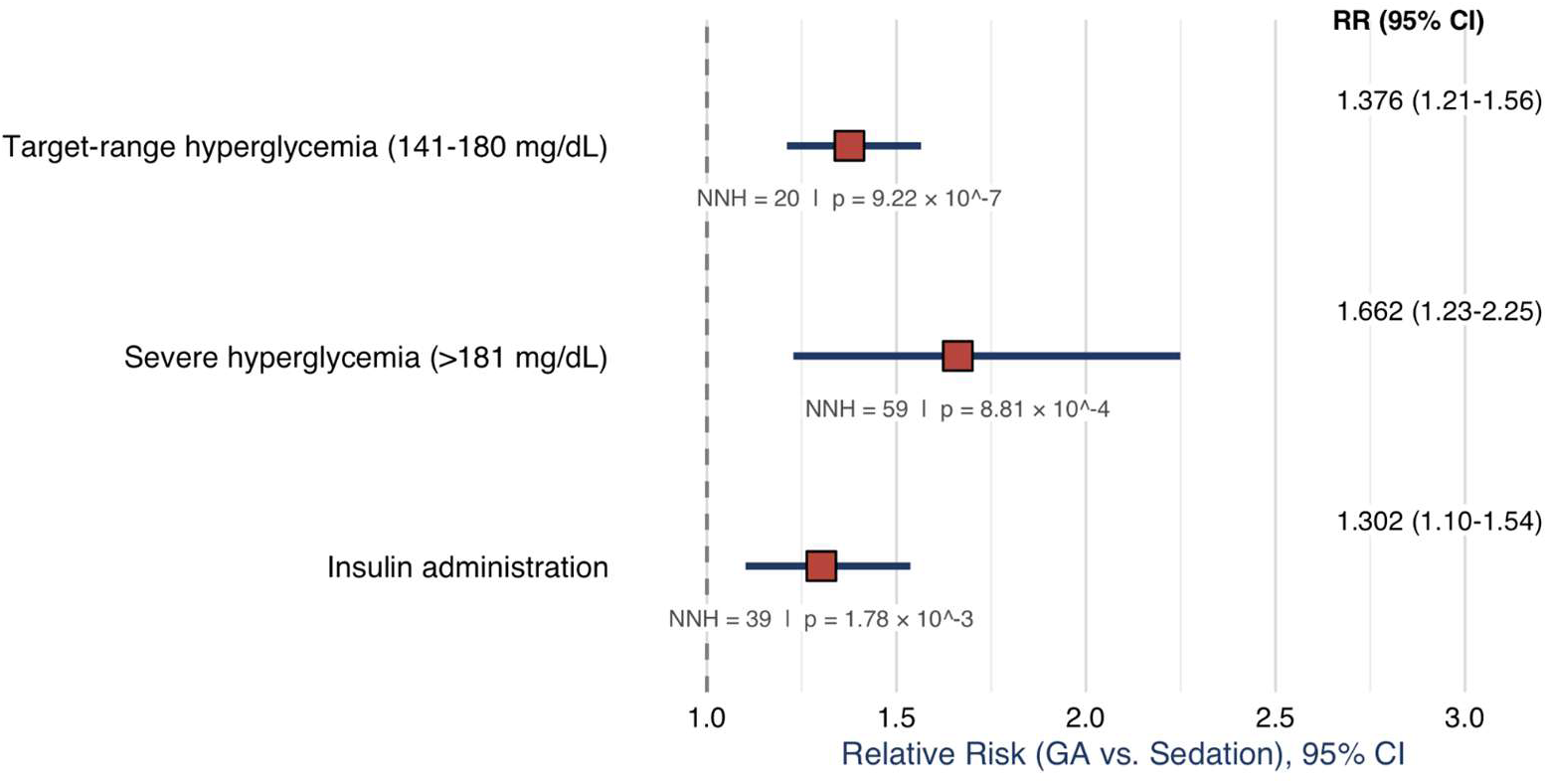
Forest Plot of Day 1 primary Glycemic outcomes Relative Risk (RR) of insulin administration, severe hyperglycemia (>181 mg/dL), and target range hyperglycemia (141-180 mg/dL) in patients managed under general anesthesia versus procedural sedation on Day 1. Squares represent point RR estimates; horizontal bars illustrate 95% confidence intervals; the dashed vertical line marks the null value (RR=1.0). All three outcomes favored increased risk under GA and survived Bonferroni Correction. NNH: number needed to harm.

**Figure 7.**
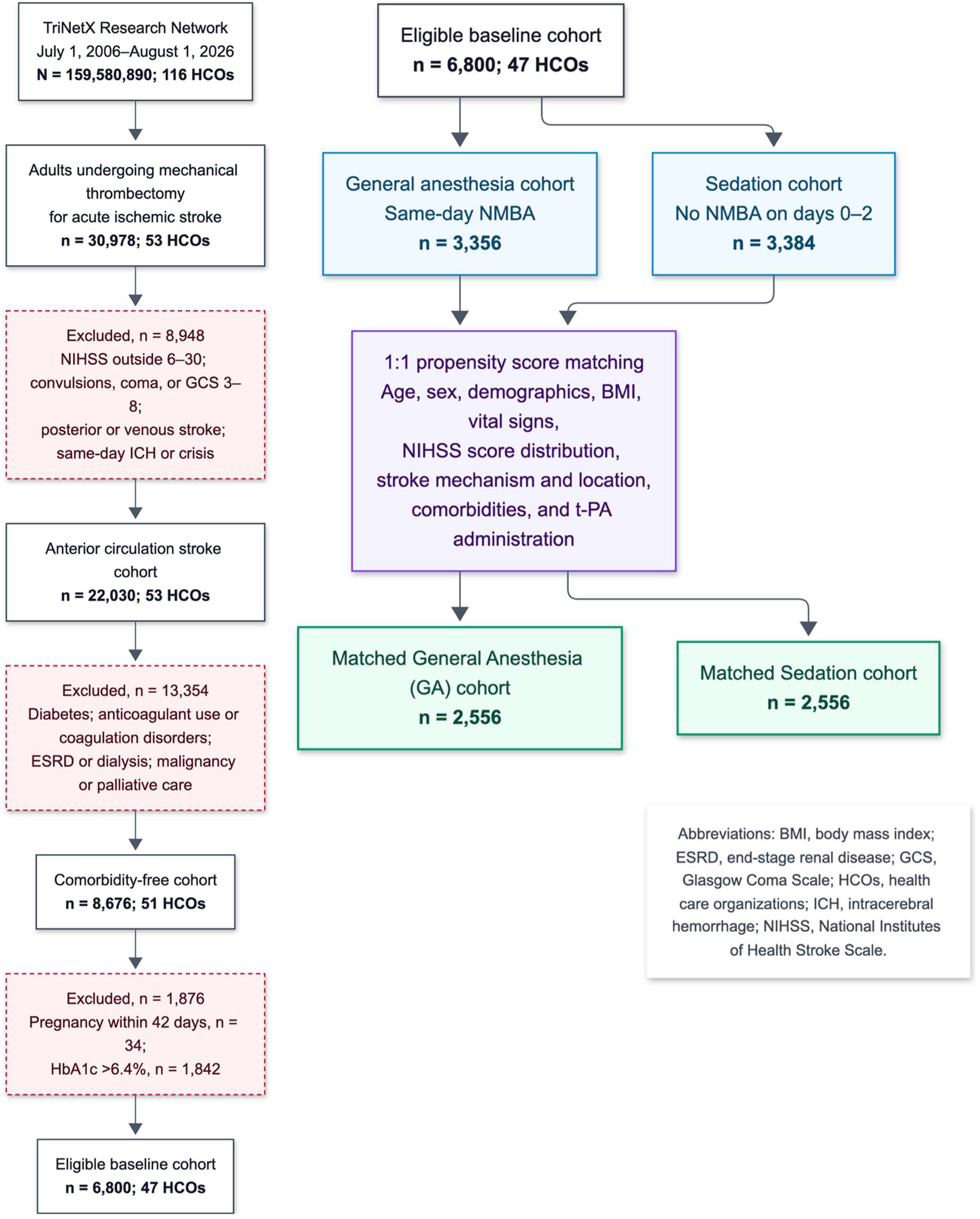
Cohort assembly for the comparison of general anesthesia (GA) and procedural sedation during mechanical thrombectomy for acute ischemic stroke.

#### Perioperative Hypoglycemia (≤ 60 mg/dL) risk analysis

The overall incidence of acute hypoglycemia (≤ 60 mg/dL) was negligible across the study population. **Day 0:** (0.43% in GA vs. 0.51% in Sedation) demonstrating no significant variance between groups (RD = –0.08%, 95% CI – 0.45%– –0.30%; z = –0.41, p = 0.682; RR = 0.846, 95% CI 0.379 – 1.889). **Day 1:** the queried outcome returned a null finding.

Quantitative sensitivity analyses (E-Values), absolute risk differences, and numbers needed to harm (NNH) for these primary outcomes are detailed in Table 2; all major findings survived Bonferroni correction.

**Table 2.** E-Value and Number Needed to Harm (NNH) summary for primary outcomes.

| Outcomes | RR | E-Value<br>(RR) | E-Value<br>(LB) | ARD | NNH | p-value |
| --- | --- | --- | --- | --- | --- | --- |
| **Day 0 Outcomes** |  |  |  |  |  |  |
| Day 0 (141-180 mg/dL) | 1.464 | 2.29 | 2.00 | 0.098 | 11 | $1.29 \times 10^{-15}$ |
| Day 0 $\geq 181$ mg/dL | 1.832 | 3.07 | 2.27 | 0.035 | 29 | $1.35 \times 10^{-7}$ |
| Day 0 Insulin administration | 1.231 | 1.764 | 1.43 | 0.04 | 25 | $2.96 \times 10^{-4}$ |
| **Day 1 Outcomes** |  |  |  |  |  |  |
| Day 1 (141-180 mg/dL) | 1.376 | 2.10 | 1.72 | 0.05 | 20 | $9.22 \times 10^{-7}$ |
| Day 1 $\geq 181$ mg/dL | 1.662 | 2.71 | 1.76 | 0.017 | 59 | $8.81 \times 10^{-4}$ |
| Day 1 Insulin administration | 1.302 | 1.93 | 1.44 | 0.026 | 39 | 0.00178 |

RR: Risk Ratio; LB: 95% CI Lower Bound of RR; ARD: Absolute Risk Difference; NNH: Number Needed to Harm
Bonferroni correction was applied to the following Day 0 and Day 1 outcomes: Target-range hyperglycemia (141-180 mg/dL), Severe Hyperglycemia ( $\geq 181$ mg/dL), and insulin administration. For 6 outcomes, $\alpha = 0.0083$ (given by $\alpha = 0.05/6$ ). All 6 outcomes survived correction.

### Surveillance Outcomes

#### Bedside POC Glucose Check

Bedside POC glucose checks were analyzed to compare parity across both arms. **Day 0:** the 33.8% (865/2,556) of the GA cohort and 33.14% (847/2,556) of the Sedation cohort received bedside POC glucose checks with a non-significant difference between groups (RD = 0.007042, 95% CI –0.018832 – 0.03292, z = 0.5334, p = 0.5937). **Day 1:** Overall testing rates were greater in the GA arm, 20.5% (523/2,556) of the GA cohort and 18.1% (462/2,556) of the Sedation cohort received bedside POC glucose checks with a significant difference between groups (RD = 2.39%, 95% CI 0.251%– 4.55%, z = 2.163, p = 0.031).

#### Laboratory Quantitative Glucose Test risk analysis

Post-procedural quantitative laboratory glucose-testing demonstrated statistical parity between treatment cohorts. **Day 0:** In the GA cohort, 10.92% of patients (279/2,556) underwent quantitative laboratory testing on Day 0 compared to 12.52% of patients (320/2,556) in the Sedation cohort. This small numerical variance was not statistically significant (RD = –1.60%, 95% CI: –3.37% – 0.167; z = –1.78, p = 0.075; RR = 0.872, 95% CI: 0.750 – 1.1014; OR =0.856, 95% CI: 0.722 – 1.016). **Day 1:** In the GA cohort, 5.83% of patients (149/2,556) underwent quantitative laboratory testing on Day 0 compared to 6.53% of patients (167/2,556) in the Sedation cohort. This small numerical variance was not statistically significant (RD = –.704%, 95% CI: –2.02% – 0.616%; z = –1.045, p = 0.296; RR = 0.892, 95% CI: 0.720 – 1.105; OR =0.886, 95% CI: 0.705 – 1.112).

#### Composite Glycemic Monitoring Parity risk analysis

To evaluate potential surveillance bias, a composite outcome of any glycemic monitoring (combining point-of-care bedside glucose checks, quantitative laboratory testing, and comprehensive metabolic panels) was assessed across both arms. **Day 0:** 57.24% (1,463/2,556) of GA patients and 56.73% (1,450/2,556) of Sedation patients undergoing post-procedural glucose monitoring (RD = 0.51%; 95% CI: –2.21%-3.22%; z = 0.37, p = 0.713; RR = 1.009, 95% CI 0.962 – 1.058; OR = 1.021, 95% CI 0.914 – 1.141). **Day 1:** 30.59% (782/2,556) of GA patients and 28.44% (727/2,556) of Sedation patients undergoing post-procedural glucose monitoring (RD = 2.15%; 95% CI: –0.35%-4.65%; z = 1.686, p = 0.0917; RR = 1.076, 95% CI 0.988 – 1.171; OR = 1.11, 95% CI 0.983 – 1.25).

## Discussion

### Methodological Rationale

We conducted a retrospective, propensity score-matched cohort study using the TriNetX Research Collaborative network to evaluate adult patients undergoing endovascular thrombectomy for anterior circulation acute ischemic stroke. NMBA administration was selected as a surrogate for anesthetic exposure as it is only administered to facilitate intubation as an adjunctive for general anesthesia, per ASA practice guidelines.^2,9^ Inclusion and exclusion criteria were modeled from existing landmark stroke trial literature.^2,5–7,9–15^ We evaluated a strictly non-diabetic cohort by excluding all patients with a prior diagnosis of diabetes mellitus or a baseline HbA1c > 6.4%.^27^ Patients were categorized into GA or Sedation cohorts based on the administration of NMBAs. Following the exclusion of patients with catastrophic baselines or delayed crossover to GA; confounding by indication was addressed via meticulous propensity score matching. Post-procedural metabolic endpoints were then evaluated across mutually exclusive glycemic risk strata alongside exogenous insulin initiation; surveillance data was queried to account for possible surveillance bias (full methodological rationale, code definitions, and cohort construction logic are detailed in Supplemental Methods 2).

### Glucose Outcomes

The present findings introduce a complementary and potentially synergistic mechanism: general anesthesia is strongly associated with a pronounced and sustained hyperglycemic response that persists well beyond the procedural period and survived rigorous sensitivity testing.^31^ The full glycemic profile reveals a coherent and internally consistent pattern of GA-associated metabolic perturbation across multiple strata of hyperglycemic severity with several structural features of the study design biasing conservatively toward the null. At the population level, mean glucose divergence was modest in absolute terms (Day 0: Δ = 4.13 mg/dL; Day 1: Δ = 4.44 mg/dL) yet achieved statistical significance (Day 0: p = 7.82 x 10^-11^, Day 1: p = 8.16 x 10^-14^). While a ∼4 mg/dL shift in mean glucose may appear clinically trivial at the individual level, this population-level displacement has important consequences: it shifts the right tail of the glucose distribution, increasing the proportion of patients who cross clinically meaningful hyperglycemic thresholds. This phenomenon is precisely what the stratified risk analyses demonstrate: target-range hyperglycemia (141-180 mg/dL) and severe hyperglycemia (≥181 mg/dL) reveal an ascending risk gradient that is more consistent with a causal signal than uniform confounding. At the target-range (141-180 mg/dL), GA conferred 46.4% relative risk increase on day 0 (RR = 1.464), escalating to 83.2% at the severe range (≥181 mg/dL; RR = 1.832). Both tiers demonstrated attenuated but persistent effects on day 1 (RR = 1.376 and 1.66; respectively).

The clinical gravity of the ≥181 mg/dL threshold becomes apparent when translated into the stress hyperglycemia ratio framework. All patients in this study were confirmed non-diabetic, with HbA1c values capped at 6.4% and a cohort median of 5.6%. A blood glucose of 181 mg/dL therefore corresponds to an SHR of at least 1.32 at the maximum eligible HbA1c; and approximately 1.59 at the cohort median, placing these patients squarely within the range associated with the most severe complications following mechanical thrombectomy. This value directly intersects the empirically derived SHR cutoff of 1.303 identified by Peng et al. in the LVO stroke population; a cutoff above which patients demonstrated significantly reduced odds of favorable 90-day functional outcome (adjusted OR 0.44; 95% CI 0.28-0.69; p<0.001).^32^ Yu et al. corroborate SHR 1.3 as a critical inflection point, above which odds of moderate-to-severe stroke severity increase sharply (OR 6.11, 95% CI: 2.64-14.15, p <0.0001).^14^ Deng et al. further demonstrated that each 0.1-point SHR increment independently increases the risk of moderate-to-severe cerebral edema by 39% (OR 1.39, 95% CI: 1.24-1.57, p <0.001).^33^ Such that the distance from an SHR to the present cohort’s threshold represents a clinically consequential accumulation of risk. Critically the SHR-outcome relationship is amplified in non-diabetic patients: subgroup analysis has demonstrated a significantly stronger association between SHR and early neurological deterioration in the non-diabetic subgroup (interaction p = 0.0033), with each unit SHR increase conferring a 6.19-fold increased risk of deterioration (95% CI 2.68-14.28) beyond an inflection point of approximately 1.06.^25^

### Insulin Administration in the GA arm

GA patients received significantly more exogenous insulin on both day 0 (RR = 1.231) and day 1 (RR = 1.30). this serves as independent clinical validation; bedside clinicians, blinded to the retrospective study hypothesis, identified and treated more hyperglycemia in GA patients. More critically, because the study captured the ‘most recent’ glucose value on each day, GA patients who received corrective insulin had their captured values pharmacologically suppressed. The observed glucose divergence therefore represents the residual metabolic difference after differential insulin correction; not pre-treatment divergence. This conservative structural bias is compounded by the study’s temporal design, by designating day 0 as the index procedural day and capturing the last glucose value rather than the peak value, the analysis preferentially captures the nadir of post-intervention glucose trajectory on the day of greatest metabolic perturbation. This design choice was deliberate, the analytic framework was constructed to ensure that any observed signal would represent a lower bound of the true effect.^23,24^ Hypoglycemia (≤60 mg/dL) was negligible in both arms on day 0 (GA: 0.43%, Sedation: 0.51%; p = 0.682) and null on day 1, confirming a wide therapeutic window for any future glucose-directed intervention in the GA population.

### Surveillance Parity

Composite glycemic monitoring rates were virtually identical with insignificant differences on both day 0 and day 1, excluding detection bias as an explanation for the observed divergence. The slight numerical trend in laboratory testing favored the sedation arm, meaning any residual surveillance asymmetry would bias against the observed effect. A small day 1 increase in bedside POC checks in the GA arm (20.5% vs 18.1%; p = 0.031) is most parsimoniously interpreted as a clinical response to already-elevated glucose values; a consequence of the exposure, not a confounder.

### Implications

No NIHSS score in the studied range (6-30) mandates a specific anesthetic approach; AHA/ASA guidance holds that either GA or procedural sedation is reasonable, leaving the choice to physician preference.^9,28^ GA is obligatory when airway protection, aspiration risk, agitation, or a prolonged procedure precludes cooperation, and pooled randomized data associate it with higher reperfusion (OR 1.73, 95% CrI 1.23-2.43).^34^ Procedural sedation is preferred in cooperative patients with protected airways, in the hemodynamically fragile (GA roughly quadruples intraprocedural hypotension, OR 4.28, 95% CrI 2.35-7.86), in those older than 70 years, and in minor/low-NIHSS occlusions.^34,35^

The “General Anesthesia Package” contributes to stress hyperglycemia through at least three distinct and compounding mechanisms that are largely absent or attenuated with procedural sedation: intubation associated catecholamine surge, anesthetic-induced impairment of insulin secretion and glucose utilization, and vasopressor driven exogenous catecholamine exposure.^17^

First, laryngoscopy and endotracheal intubation provoke an acute sympathoadrenal response characterized by significant elevations in circulating catecholamines. Hassan et al. demonstrated augmented plasma epinephrine concentrations following intubation compared with laryngoscopy alone, with steeper dose-response slopes and slower regression. Intubation has also been shown to increase norepinephrine plasma concentrations as well.^36,37^ These catecholamines directly stimulate hepatic glycogenolysis and gluconeogenesis while inhibiting insulin secretion, producing an immediate hyperglycemic stimulus that is entirely bypassed in patients managed with procedural sedation.

Second, the anesthetic agents themselves exert independent deleterious effects on glucose homeostasis. Volatile anesthetics; the maintenance agents commonly used during GA; exhibit well documented suppression of glucose-stimulated insulin secretion by opening adenosine triphosphate-sensitive potassium (K-ATP) channels in pancreatic β-cells, reducing ATP sensitivity of these channels and thereby uncoupling the glucose-sensing mechanism from insulin release.^38–40^ Even when GA is maintained with total intravenous anesthesia (TIVA), usually with propofol; propofol is not metabolically inert.^41^ At GA relevant doses, propofol induces marked whole-body insulin resistance, with decreased insulin-stimulated glucose uptake in skeletal muscle and heart and increased hepatic glucose output.^41,42^ Additionally, practice patterns demonstrate that 51% of US centers favor using volatile anesthetics to maintain patients for mechanical thrombectomy, whereas 6 of the 7 RCTs comparing GA vs procedural sedation used propofol-based TIVA as the GA maintenance agent.^35,43^ Meaning that the evidence base demonstrating GA’s benefit in thrombectomy is built almost entirely on propofol-based protocols; which may not reflect the real-world practice patterns.

Third, many anesthetics are profoundly vasodilatory and GA-induced hemodynamic instability necessitates vasopressor administration at substantially higher rates than procedural sedation, with a recent Bayesian meta-analysis demonstrating 4.3-fold higher odds of intraoperative hypotension under GA (OR: 4.28, 95% CrI 2.35-7.86).^34^ The vasopressors commonly used to counteract this hypotension; norepinephrine, epinephrine, and phenylephrine, are exogenous catecholamines or catecholamine analogues that perpetuate hyperglycemia through the exact same metabolic pathways as endogenous stress-associated catecholamine surges.^44–47^ Norepinephrine at clinically used pressor doses reduces insulin sensitivity, and catecholamines in the setting of critical illness aggravate hypermetabolism by promoting hyperglycemia and increasing oxygen demands.^48^ This vasopressor-driven metabolic burden represents a sustained, iatrogenic source of stress hyperglycemia that persists beyond the initial intubation stimulus and compounds the effects of the anesthetic agents themselves.

By contrast, the sedative agents used for procedural sedation during thrombectomy; typically low-dose propofol infusions, fentanyl boluses, dexmedetomidine, or midazolam, have a fundamentally different metabolic footprint.^35,49^

Dexmedetomidine, commonly used as a sedative, is a selective α2-adrenergic agonist that inhibits central norepinephrine release, dexmedetomidine does not alter the incidence of intraoperative hyperglycemia (27.4% vs 22.5%, p = 0.167), though it modestly reduced glycemic variability.^49,50^ The benzodiazepines (midazolam and diazepam) used in procedural sedation protocols lack the direct pancreatic or hepatic glucose transport effects characteristic of either volatile agents or high-dose propofol.^41^ Thus the sedative regimens employed during procedural sedation for thrombectomy are pharmacologically less disruptive to glucose homeostasis than the anesthetic agents required for GA, independent of the hemodynamic and airway-related mechanisms discussed above.^41^

Furthermore, even when propofol is used as the sedative agent during procedural sedation for thrombectomy, the cumulative dose is substantially lower than that required for GA maintenance. Procedural sedation protocols in thrombectomy RCTs employed low-dose propofol infusions supplemented with fentanyl boluses, whereas GA protocols required propofol at hypnotic doses sufficient to abolish consciousness and suppress airway reflexes throughout the procedure.^35^ This dose differential is clinically relevant because propofol’s metabolic effects; including systemic whole-body insulin resistance, follows a dose-response curve.^42^ Patients receiving low-dose propofol for procedural sedation would be expected to experience attenuated glucose derangements compared with those receiving continuous high-dose propofol infusions for GA-maintenance. Additionally, the shorter procedural sedation exposure window where propofol is titrated intermittently, rather than infused continuously; further limits total drug load, reducing cumulative metabolic insult.

### Limitations

A recognized limitation of administrative-database extraction is that reimbursement-linked endpoints and lab values (mortality, procedure codes, glucose labs) are captured reliably, whereas patient-centered functional status; most notably the 90-day modified Rankin-scale (mRs), is not encoded in structured ICD-10-CM fields and can be recovered only through labor-intensive chart review (present study, n = 5,112).^51^ This limitation is mitigated by our choice of endpoints, severe range hyperglycemia (≥ 181 mg/dL) which has been well established in the literature as an independent predictor of poor 90-day functional outcome, as well as morbidity and mortality; and thus, function as validated surrogates for the long-term disability that structured coding cannot capture.^4,13–15,33,52,53^

Although cohorts were 1:1 matched on baseline stroke severity (NIHSS 6-30), stroke mechanism, and occlusion location, the clinical determinants that most directly drive selection of general anesthesia; depressed level of consciousness, agitation precluding awake sedation, and airway compromise, were not captured. These variables cannot be assumed glycemically neutral. Stress hyperglycemia arises from hypothalamic-pituitary-adrenal axis activation and sympathoadrenal outflow, producing glucocorticoid and catecholamine-driven glycogenolysis, gluconeogenesis, and insulin resistance; agitation, respiratory distress, and the systemic arousal accompanying large hemispheric infarction are themselves expressions of that same pathway, and impaired consciousness has been directly associated with stress hyperglycemia in non-diabetic stroke cohorts.^54,55^ Additionally, core volume, collateral grade, and hypoperfusion intensity ratio were unavailable in the present dataset and remain the principal residual confounding domain. Residual confounding by indication, specifically uncaptured stroke severity (procedural complexity, poor collateral vessel status, extensive ischemic core, etc.) could theoretically influence both the clinical decision to utilize general anesthesia and trigger a heightened endogenous neuroendocrine stress response.

Our argument is therefore one of magnitude rather than absence: to fully explain the observed association, an unmeasured confounder would need to be associated with both anesthetic modality and severe day 0 hyperglycemia by a risk ratio of at least 3.07 (2.27 to render the lower confidence bound non-significant) above and beyond the measured covariates. Additionally for the severe day 1 hyperglycemia outcome, representing at least 24 hours post-procedural sustained hyperglycemia; an unmeasured confounder would need to be associated with anesthetic modality and severe hyperglycemia by a risk ratio of 2.71 (1.76 to render the lower confidence bound non-significant). This confounder would need to increase the risk after adjusting for all matched covariates and insulin regimens. While unmeasured confounders (e.g., collateral failure) certainly drives metabolic stress, it is clinically unlikely that uncaptured severity alone exerts a >127%-76% risk surge on anesthetic selection without manifesting in captured baseline variables (such as explicit NIHSS diagnoses, baseline GCS, or immediate post-procedure hemorrhagic transformation, etc.), which were balanced or excluded in our design.

Furthermore, factors such as procedural duration are a facet of general anesthesia, intubation and induction are known to delay the time to puncture; and cannot be considered a confounder.^56^ While E-values cannot account for biases other than unmeasured confounding and have a near-linear relationship with the effect estimate, no readily identifiable confounder in the stroke thrombectomy literature approaches this threshold after propensity matching, lending further credibility to the observed association.^31^ Differential outcome ascertainment is an unlikely driver of these findings: the present study found remarkable parity in surveillance procedures in both study arms; additionally, post-procedure surveillance and assessment follow uniform ICU and/or institutional protocols applied irrespective of anesthetic modality. ^35^

### Conclusion

As a retrospective cohort study, the present analysis demonstrates association rather than causation; however, the consistency and biological plausibility of the observed relationship between anesthetic modality and perioperative glycemic derangement warrant prospective investigation. Nevertheless, these findings highlight a significant knowledge gap. Current neurointerventional guidelines leave anesthetic modality to physician preference without acknowledging differential metabolic consequences.^26,28^ The recommended glucose target of 140-180 mg/dL derives from evidence that did not isolate non-diabetic patients: the SHINE trial enrolled an approximately 80% diabetic population, found no benefit from intensive control (80-130 mg/dL), and the 2026 AHA/ASA guidelines now recommend against intensive targets due to hypoglycemia risk.^16,26,28,57^ Whether a population-specific target for non-diabetic patients experiencing anesthetic-driven stress hyperglycemia might improve outcomes remains entirely unanswered.

The contrast with cardiac surgery is striking. CABG; a procedure involving reperfusion of a glycemically sensitive, ischemia-vulnerable organ, has benefited from decades of dedicated research, culminating in Class I guideline recommendations for intraoperative insulin infusions and formal quality metrics tracking glycemic control at the facility level.^21,22^ Yet the brain is arguably more vulnerable: it has minimal energy reserves, relies almost exclusively on oxidative glucose metabolism, and depletes ATP within minutes of flow cessation.^20^ Hyperglycemia during focal cerebral ischemia directly expands infarct volume through enhanced tissue acidosis and impaired reperfusion recovery; mechanisms with no direct parallel in cardiac surgery, where glycemic harm is primarily infectious and arrhythmic.^18,19,21,58^ Despite this heightened vulnerability, no guideline addresses glycemic management as a function of anesthetic technique during thrombectomy, no quality metric for perioperative glycemic control exists, and no trial has evaluated glycemic targets in non-diabetic stroke patients undergoing endovascular reperfusion. The present study suggests that anesthetic modality may be an underappreciated and modifiable driver of perioperative hyperglycemia warranting greater scrutiny.

## Data Availability

The data that support the findings of this study were obtained from the TriNetX Research Network (TriNetX LLC) under a license/data use agreement and are not publicly available. Access to TriNetX data is available to qualified researchers through TriNetX (https://trinetx.com) upon reasonable request and completion of the required data use agreement. The authors do not have permission to share the underlying patient-level data. The analytic methods, cohort definitions, diagnosis/procedure codes, and statistical code used in this study are available from the corresponding author upon reasonable request.

## Acknowledgements

The authors acknowledge the TriNetX Research Federation for providing access to the de-identified electronic health record data used in this study.

## Supplemental Material

Supplemental Methods

1. Code Definitions
2. Methodological Rationale
3. Raw Query Architecture

References 2-30

